# COVID-19 Mortality in Women and Men in Sub-Saharan Africa: A Cross Sectional Study

**DOI:** 10.1101/2021.07.31.21261422

**Authors:** Jyoti Dalal, Isotta Triulzi, Ananthu James, Benedict Nguimbis, Gabriela Guizzo Dri, Akarsh Venkatasubramanian, Noubi Tchoupopnou Royd Lucie, Sara Botero Mesa, Claire Somerville, Giuseppe Turchetti, Beat Stoll, Jessica Lee Abbate, Franck Mboussou, Benido Impouma, Olivia Keiser, Flávio Codeço Coelho

## Abstract

**Objective:** To investigate differences of COVID-19 related mortality among women and men across sub-Saharan Africa (SSA) from the beginning of the pandemic.

**Design:** A cross sectional study.

**Setting:** Data from 20 member nations of the WHO African region until September 1, 2020.

**Participants:** 69,580 cases of COVID-19, stratified by sex (men, n=43071; women, n=26509) and age (0-39 years, n=41682; 40-59 years, n=20757; 60+ years, n=7141).

**Main outcome measures:** We computed the SSA- and country-specific case fatality rates (CFRs) and sex-specific CFR differences across various age groups, using a Bayesian approach.

**Results:** A total of 1,656 (2.4% of total cases reported; 1656/69580) deaths were reported, with men accounting for 1168/1656 (70.5%) of total deaths. In SSA, women had a lower CFR than men (mean *CFR*_*diff*_ = -0.9%; 95% credible intervals -1.1% to -0.6%). The mean CFR estimates increased with age, with the sex-specific CFR differences being significant among those aged 40 or more (40-59 age-group: mean *CFR*_*diff*_ = -0.7%; 95% credible intervals -1.1% to -0.2%; 60+ age-group: mean *CFR*_*diff*_ = -3.9%; 95% credible intervals -5.3% to -2.4%). At the country level, seven of the twenty SSA countries reported significantly lower CFRs among women than men overall. Moreover, corresponding to the age-specific datasets, significantly lower CFRs in women than men were observed in the 60+ age-group in seven countries and 40-59 age-group in one country.

**Conclusions:** >Sex and age are important predictors of COVID-19 mortality. Countries should prioritize the collection and use of sex-disaggregated data to understand the evolution of the pandemic. This is essential to design public health interventions and ensure that policies promote a gender sensitive public health response.

**Summary Box:** *What is already known on this topic:*

- Little is known on the impact of COVID-19 among different sexes and age-groups in sub-Saharan Africa (SSA).
- The availability of data on COVID-19 cases and deaths, disaggregated by both age and sex from the WHO African region has been scarce.
- In most of the non-African countries, sex-specific COVID-19 severity and mortality were substantially worse for men than for women, during the first wave of the novel coronavirus (COVID-19) pandemic.

*What this study adds:*

- To the best of our knowledge, this is the largest study focussing on the COVID-19 related fatalities among men and women in SSA, and it confirmed that both sex and age are important predictors of COVID-19 mortality in SSA, similar to other regions.
- In SSA, overall, men had a higher case fatality rate (CFR) than women. When disaggregated by age, this difference persisted only in individuals aged 40 or more. 7 among the 20 SSA countries included in this study also reported significantly higher CFRs in men than women for the age-aggregated dataset.
- Public health prevention activities and responses should take into account gender differences in terms of disease severity and mortality, especially among men aged 40 or more in SSA.

## Introduction

Severe acute respiratory syndrome coronavirus 2 (SARS-CoV-2), first reported in December 2019, rapidly underwent an exponential increase in cases and related fatalities, affecting almost every country in the world. Globally, COVID-19 had approximately infected 25,000,000 individuals and caused 840,000 deaths, resulting in a case fatality rate (CFR) of around 3.8% as of August 31, 2020. At that time, Africa (representing ∼17% of the world’s population) accounted for 4% of total confirmed cases and 3% of the total deaths reported, resulting in a CFR of around 2.1%, much lower than the global estimate ^1^, making it one of the least affected continents ^2^. The availability of data on COVID-19 cases and deaths that are disaggregated by both age and sex from the World Health Organization (WHO) African region has been scarce. As per the report published by Global Health 5050 in Jan 2021, the total number of COVID-19 cases disaggregated by sex in the region accounted for merely 2.5% of total cases and 2.7% of total deaths reported globally ^3^. Moreover, among the countries with sex information on cases, only 36% shared age information, whereas among the countries with sex information on deaths, only 42% shared age-disaggregated data ^4^.

Increasing evidence has demonstrated that infections caused by SARS-CoV-2 lead to major variations in disease severity and mortality between women and men ^5^. During the first wave of the novel coronavirus (COVID-19) pandemic in the United States, China, Singapore, South Korea, and multiple European countries, sex-specific COVID-19 susceptibility, severity, and mortality were substantially worse for men than for women ^6^. Furthermore, globally, men demonstrate increased mortality due to SARS-CoV-2 infection compared to women ^7,8^. The difference in the fatality of COVID-19 could be attributed to sex-based biological, and/or gender factors (social, behavioral, or lifestyle). However, few policies and public health responses to COVID-19, take gender and sex into account ^9,10^, although both played considerable roles in transmission, course and outcome of infectious diseases, such as Ebola, Dengue and Severe acute respiratory syndrome (SARS). For this reason, sex- and age-disaggregated data are essential to understand the course of the pandemic, identify existing or arising gender-related health inequities and the subsequent formations of gender-sensitive responses to the pandemic ^11^.

Since there is little published research on the impact of COVID-19 among different sexes and age-groups in sub-Saharan Africa (SSA), our objective was to estimate COVID-19 mortality rates, as determined by crude CFR, among men and women within 20 SSA countries, from the beginning of the pandemic to early September 2020.

## Methods

### Study design and settings

We carried out a cross-sectional analysis of the COVID-19 cases diagnosed until September 1, 2020. Our primary data sources were national situation reports made public by all SSA countries experiencing the COVID-19 pandemic. The data from each country report were extracted and merged into a linelist. Among the 47 member states comprising the WHO African region, 35 shared sex- and age-disaggregated data with the WHO. We included countries that shared data for at least 120 days from the identification of the index case. Twenty countries, Angola, Botswana, Burkina Faso, Chad, Congo, Eswatini, Gambia, Guinea, Kenya, Liberia, Mauritius, Mozambique, Namibia, Niger, Rwanda, São Tomé e Principe, Sierra Leone, Senegal, Seychelles, and Uganda, met the criteria and were included (Figure 1). Among these 20, we only included the cases with confirmed information about sex, age, patient’s outcome status, and date of reporting. The time period covered for each country is shown in Table 1.

**Table 1:**
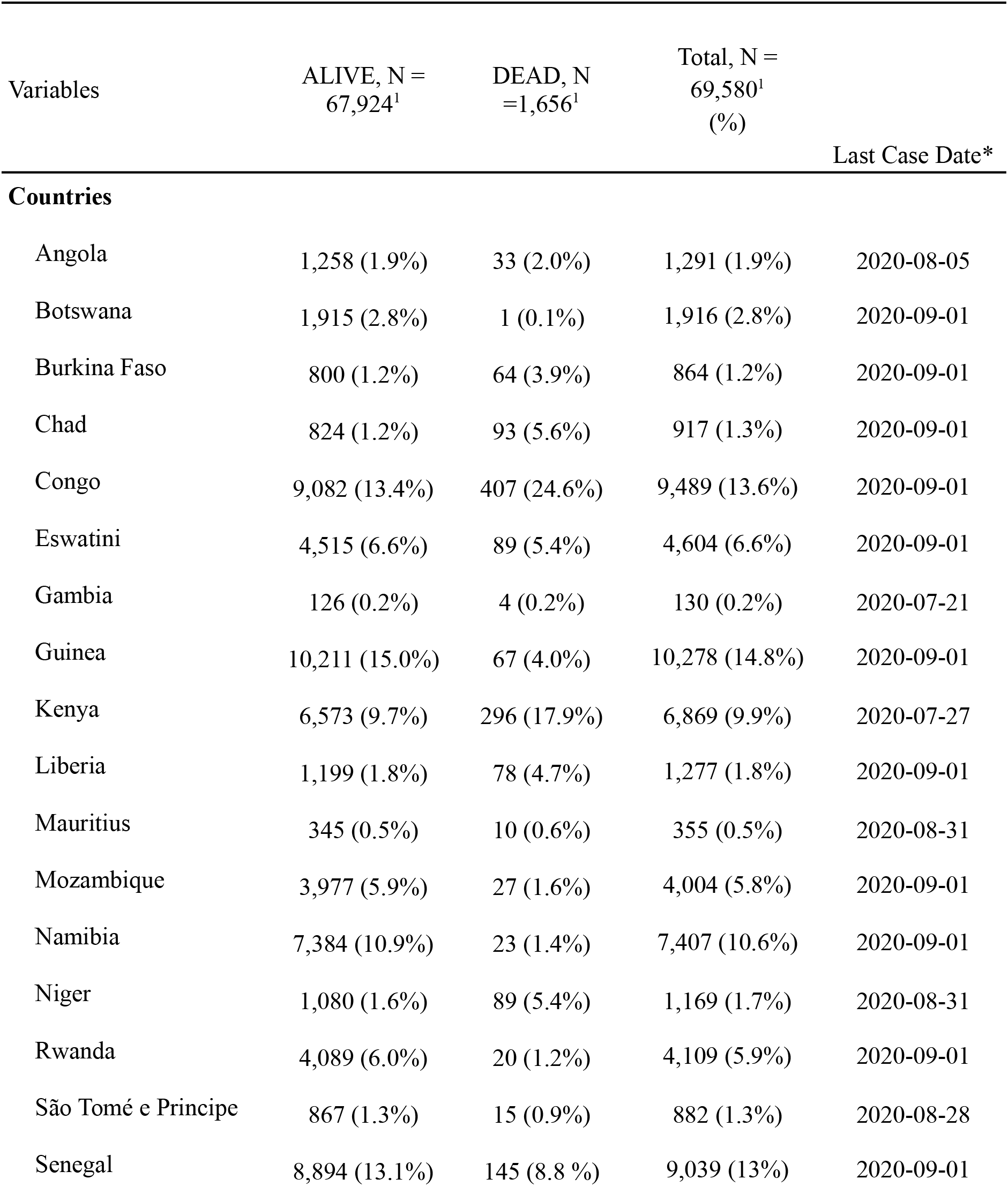
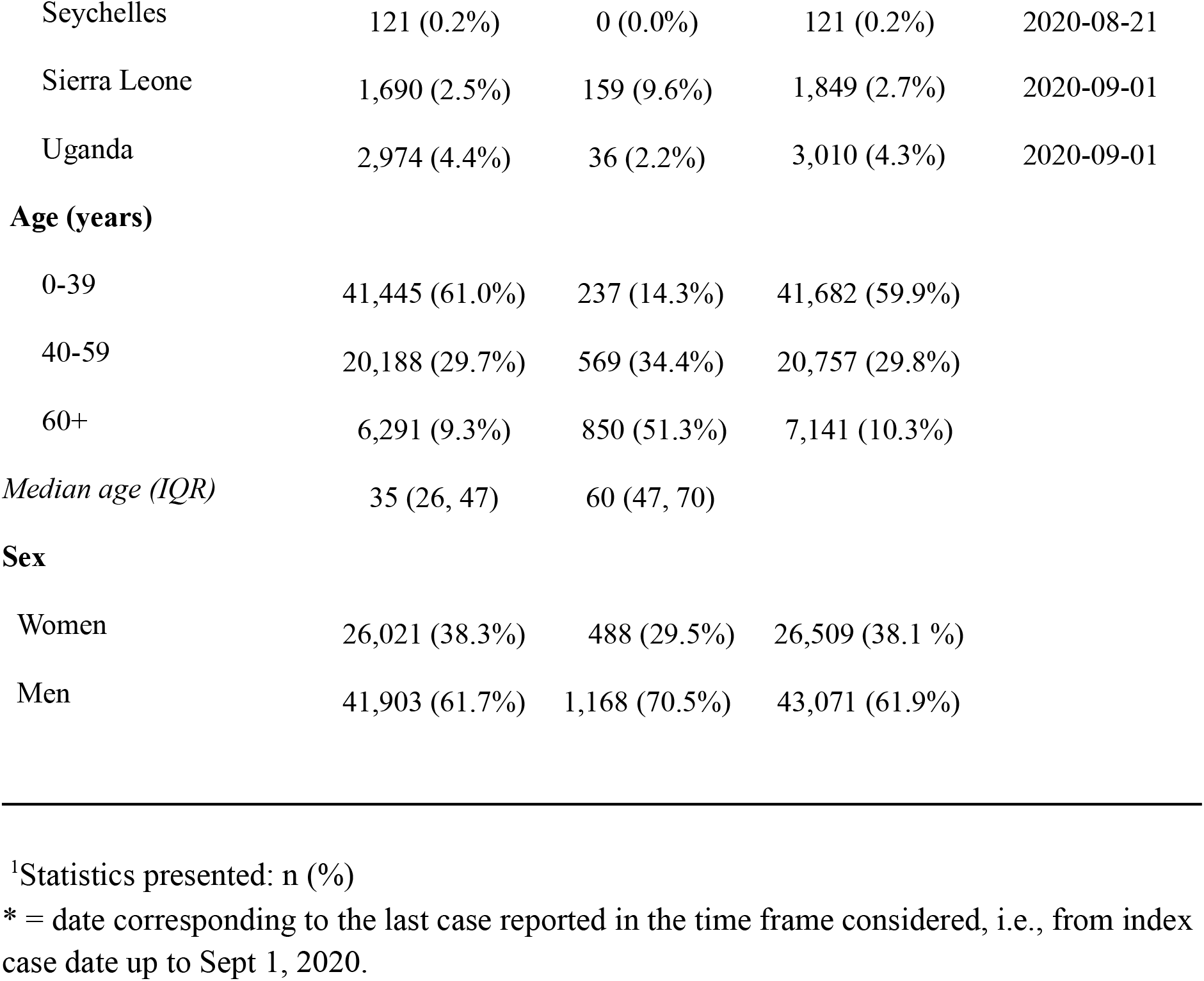
Sample characteristics of the data included in this study. Total (= 69580) is the sum of cases with ALIVE (= 67,924) and DEAD (= 1,656) clinical outcomes, at the time of data collection, Sept 1, 2020. IQR: Interquartile range.

| Variables | ALIVE, N =<br>67,924<sup>1</sup> | DEAD, N<br>=1,656<sup>1</sup> | Total, N =<br>69,580<sup>1</sup><br>(%) | Last Case Date* |
| --- | --- | --- | --- | --- |
| **Countries** |  |  |  |  |
| Angola | 1,258 (1.9%) | 33 (2.0%) | 1,291 (1.9%) | 2020-08-05 |
| Botswana | 1,915 (2.8%) | 1 (0.1%) | 1,916 (2.8%) | 2020-09-01 |
| Burkina Faso | 800 (1.2%) | 64 (3.9%) | 864 (1.2%) | 2020-09-01 |
| Chad | 824 (1.2%) | 93 (5.6%) | 917 (1.3%) | 2020-09-01 |
| Congo | 9,082 (13.4%) | 407 (24.6%) | 9,489 (13.6%) | 2020-09-01 |
| Eswatini | 4,515 (6.6%) | 89 (5.4%) | 4,604 (6.6%) | 2020-09-01 |
| Gambia | 126 (0.2%) | 4 (0.2%) | 130 (0.2%) | 2020-07-21 |
| Guinea | 10,211 (15.0%) | 67 (4.0%) | 10,278 (14.8%) | 2020-09-01 |
| Kenya | 6,573 (9.7%) | 296 (17.9%) | 6,869 (9.9%) | 2020-07-27 |
| Liberia | 1,199 (1.8%) | 78 (4.7%) | 1,277 (1.8%) | 2020-09-01 |
| Mauritius | 345 (0.5%) | 10 (0.6%) | 355 (0.5%) | 2020-08-31 |
| Mozambique | 3,977 (5.9%) | 27 (1.6%) | 4,004 (5.8%) | 2020-09-01 |
| Namibia | 7,384 (10.9%) | 23 (1.4%) | 7,407 (10.6%) | 2020-09-01 |
| Niger | 1,080 (1.6%) | 89 (5.4%) | 1,169 (1.7%) | 2020-08-31 |
| Rwanda | 4,089 (6.0%) | 20 (1.2%) | 4,109 (5.9%) | 2020-09-01 |
| São Tomé e Príncipe | 867 (1.3%) | 15 (0.9%) | 882 (1.3%) | 2020-08-28 |
| Senegal | 8,894 (13.1%) | 145 (8.8 %) | 9,039 (13%) | 2020-09-01 |
| Seychelles | 121 (0.2%) | 0 (0.0%) | 121 (0.2%) | 2020-08-21 |
| Sierra Leone | 1,690 (2.5%) | 159 (9.6%) | 1,849 (2.7%) | 2020-09-01 |
| Uganda | 2,974 (4.4%) | 36 (2.2%) | 3,010 (4.3%) | 2020-09-01 |
| **Age (years)** |  |  |  |  |
| 0-39 | 41,445 (61.0%) | 237 (14.3%) | 41,682 (59.9%) |  |
| 40-59 | 20,188 (29.7%) | 569 (34.4%) | 20,757 (29.8%) |  |
| 60+ | 6,291 (9.3%) | 850 (51.3%) | 7,141 (10.3%) |  |
| *Median age (IQR)* | 35 (26, 47) | 60 (47, 70) |  |  |
| **Sex** |  |  |  |  |
| Women | 26,021 (38.3%) | 488 (29.5%) | 26,509 (38.1 %) |  |
| Men | 41,903 (61.7%) | 1,168 (70.5%) | 43,071 (61.9%) |  |

<sup>1</sup>Statistics presented: n (%)
\* = date corresponding to the last case reported in the time frame considered, i.e., from index case date up to Sept 1, 2020.

**Figure 1.**
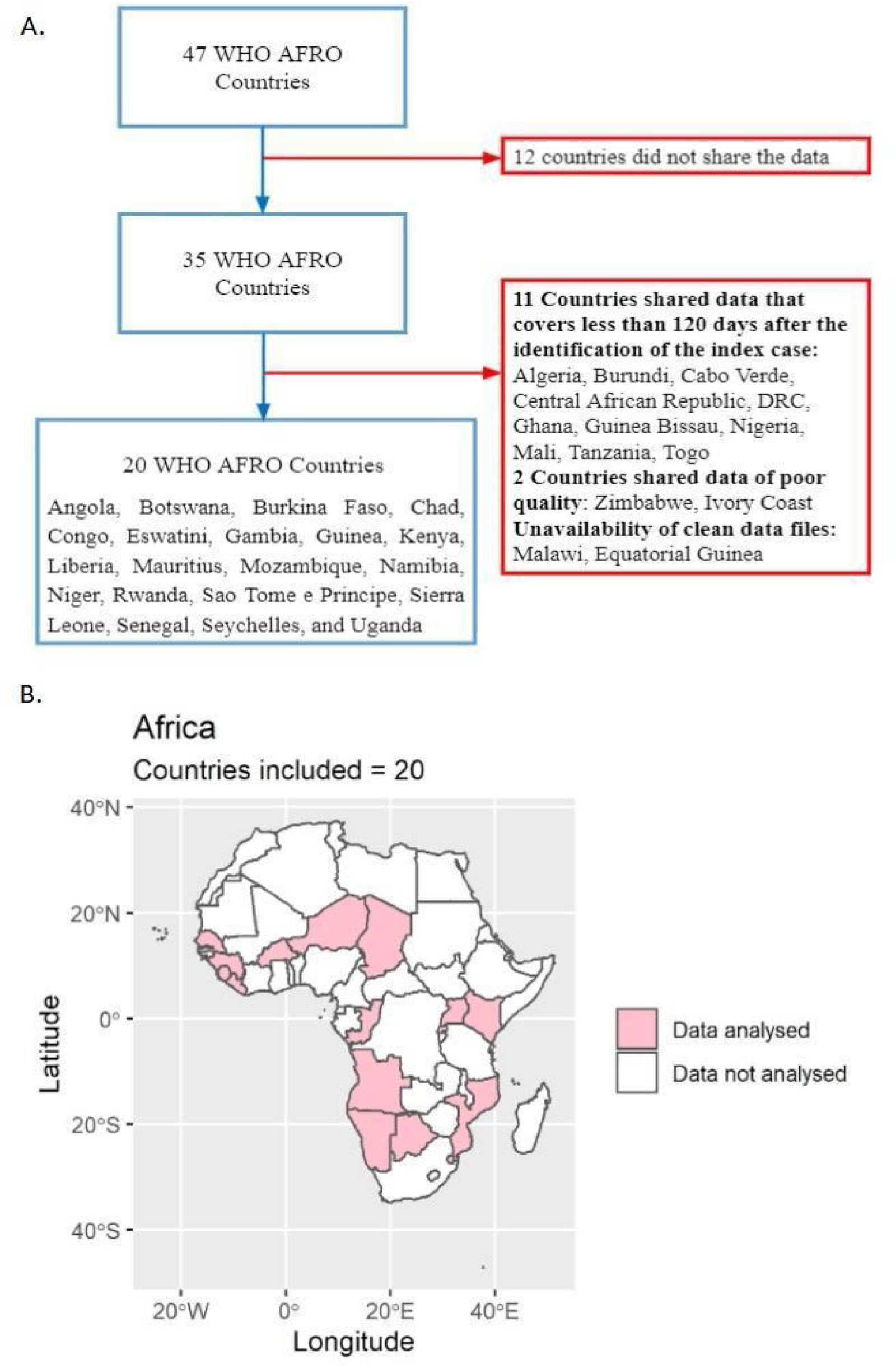
A) Flowchart illustrating the inclusion and exclusion criteria used to select countries; B) Map showing the 20 WHO AFRO countries included in the final analysis.

### Variables

For each country, we considered sex (women and men), age, date of reporting, and clinical outcomes. We classified the cases into three age categories: 0-39, 40-59 and 60+ years. Clinical outcomes were classified as dead or alive. The alive category encompassed patients who were still alive at the time of reporting: this included recovered cases.

### Statistical Analysis

Bayesian inference offers a robust inferential framework that accurately estimates the probability of rare events. This allows us to be explicit in our prior belief of observing no gender differences in CFR, making the detected differences even more relevant. It is important to note that we are not looking just at the case counts, but at the proportion of deaths to non-deaths. Thus, observing 0 deaths is as informative as any other number of deaths, to estimate the probability of dying. Hence, we included countries that also demonstrated a low number of deaths.

We followed a Bayesian approach to compute SSA- and country-specific CFRs, sex- and age-specific crude CFR differences, and their corresponding posterior probabilities. For priors, we used non-informative priors, as age-specific CFRs from the SSA region were not available. We modeled deaths as a binomial random variable, *Binomial(N, p)*, with *p* being the ratio of deaths to cases, and *N* being the number of confirmed cases. Thus, the parameter *p* represents the CFR, to which was attributed a non-informative *Beta(α = β = 0*.*33)* prior distribution. This particular neutral parameterization for the Beta was first proposed by Kerman^12^.

For each country, we computed the Bayesian estimates for sex- and age-specific CFRs, overall (all age-groups combined) CFRs, and the crude CFR differences *CFR*_*diff*_. A negative *CFR*_*diff*_ demonstrates that CFR was higher in men than in women.

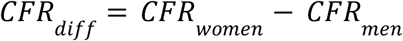

To compute the Bayesian estimates, we employed the Markov chain Monte Carlo (MCMC) sampling algorithm, No-U-Turn Sampler (NUTS) ^13^, that was implemented in the probabilistic programming (PP) package PyMC3 ^14^. To evaluate the convergence of MCMC, we generated four independent Markov chains and the resulting marginal distributions were compared using the Gelman-Rubin diagnostic (R_hat) ^15^. A strict criterion of *Rc* < 1. 1 was used to declare the convergence of the Markov chain.

For each Bayesian estimate, we reported the corresponding posterior beta distributions along with their mean and the 95% highest posterior density (HPD) intervals.

*Sensitivity Analysis:* To assess the robustness of our prior, we performed the same analysis using two more priors based on the binomial likelihood: *Beta*(1,1) or Bayes-Laplace prior ^16^ and *Beta*(0.5, 0.5) or Jeffreys prior ^17^.

## Results

### Sub-Saharan Africa estimates

#### Sample characteristics of the COVID-19 cases and deaths

As of September 1, 2020, a total of 69,580 COVID-19 cases and 1,656 (2.4% of total cases; 1656/69580) COVID-19-related deaths were reported by the 20 member states in the WHO African region included in our research (Figure 1B). Men accounted for 61.9% (43071/69580) of the total reported cases and 70.5% (1168/1656) of total deaths. 51.3% (850/1656) of the total deaths occured in the ‘60+’ age group. Among the confirmed cases of COVID-19, the median age of those still alive was 35 years, while it was 60 years for individuals who died, at the time of data collection (Table 1).

#### Mean CFR estimates in men and women

Mean CFR estimates increased with age for both men and women (Figure 2 and Table S1).

**Figure 2:**
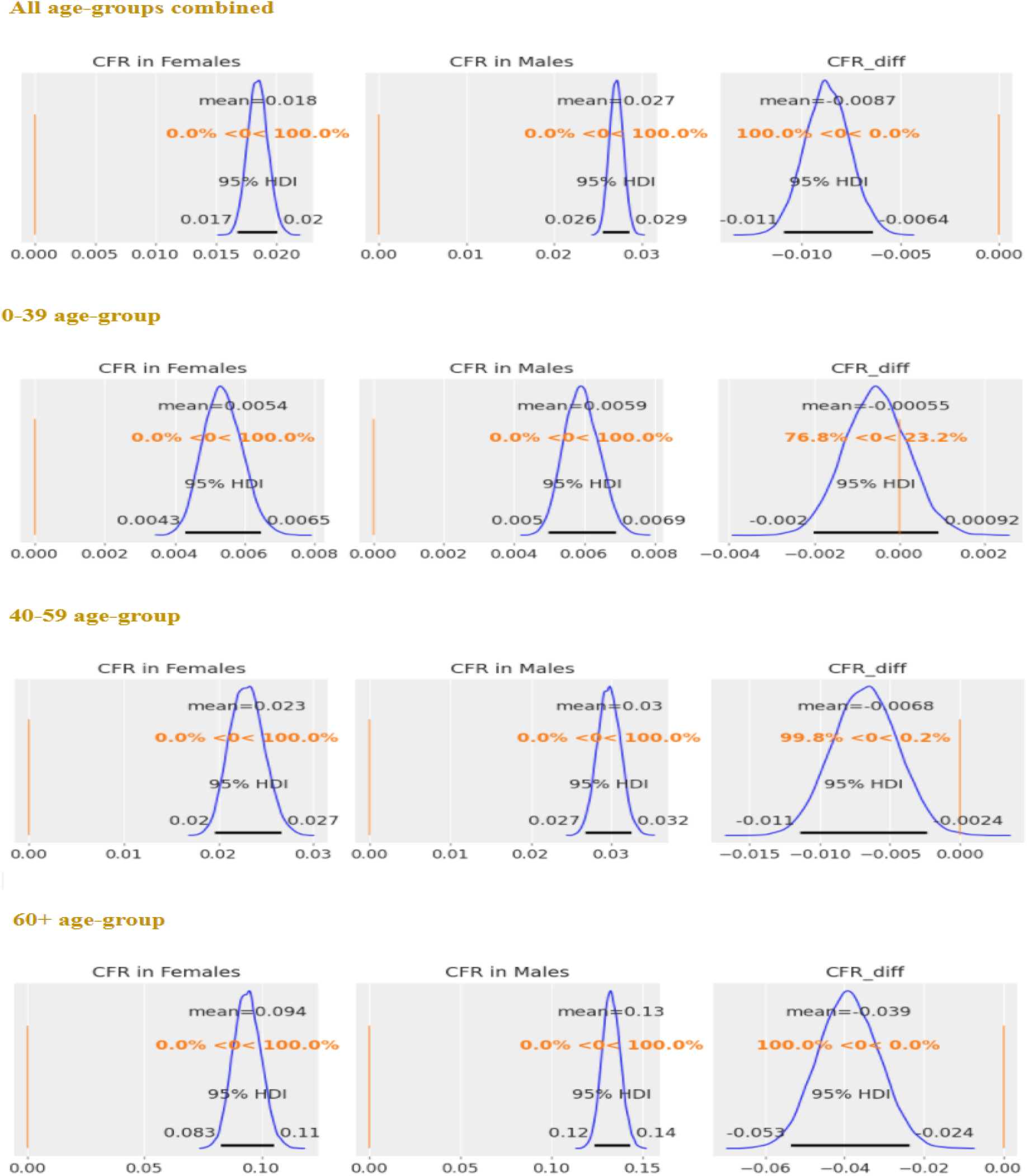
*SSA-specific estimates, after merging the data from 20 SSA countries:* posterior distributions of CFR in women, CFR in men, and *CFR*_*diff*_ (= CFR in women - CFR in men), for each age-group and all age-groups combined. The horizontal black line represents the 95% HPDs. 95% HPD intervals are the same as that of 95% HDIs (= highest density intervals). 95% HPDs for *CFR*_*diff*_ don’t contain 0 for estimates obtained in both 40-59 and 60+ age-groups and overall (all age-groups combined), indicating significant differences between sex-specific CFRs.

#### Risk differences

*CFR*_*diff*_: Overall, CFR was significantly higher in men than in women (mean *CFR*_*diff*_ = -0.9%; 95% HPD intervals -1.1% to -0.6%). When analysing the data by age, both the ‘40-59’ (mean *CFR*_*diff*_ = -0.7%; 95% HPD intervals -1.1% to -0.2%) and ‘60+’ (mean *CFR*_*diff*_ = -3.9%; 95% HPD intervals -5.3% to -2.4%) age-groups demonstrated significantly higher CFRs for men than for women. There were no significant differences between sex-specific CFRs in the ‘0-39’ age-group (mean *CFR*_*diff*_ = -0.1%; 95% HPD intervals -0.2% to 0.1%) (Figure 2 and Table S1).

### Country-specific estimates

#### Sample characteristics of the COVID-19 cases and deaths

Among the 20 countries included in this study, the countries with the highest number of cases were Guinea (14.8%, n = 10,278), Congo (13.6%, n = 9,489), Senegal (13.0%, n = 9,039), Namibia (10.6%, n = 7,407), and Kenya (9.9%, n = 6,869). Congo accounted for the largest proportion of all deaths reported (24.6%, n = 407), followed by Kenya (17.9%, n = 296; Table 1).

#### Mean CFR estimates in women and men

Crude mean CFR estimates ranged from 0.2% (0.1% to 0.4%) in Guinea to 9.2% (5.5% to 13.0%) in Chad among women, and from 0 in Botswana to 10.5% (8.3% to 12.9%) in Chad among men (Table S2). Mean crude CFR estimates increased with age, except for Angola, Mauritius, São Tomé e Principe, and Seychelles (Figure S2 & Table S2).

#### Risk differences

*CFR*_*diff*_ : After combining the data from all the age-groups, CFRs were significantly higher in men than women in seven SSA countries: Angola (mean *CFR*_*diff*_ = -1.8%; 95% HPD intervals -3.4% to -0.1%), Eswatini (mean *CFR*_*diff*_ = -1.3%; 95% HPD intervals -2.1% to -0.5%), Guinea (mean *CFR*_*diff*_ = -0.7%; 95% HPD intervals -0.9% to -0.4%), Kenya (mean *CFR*_*diff*_ = -1.4%; 95% HPD intervals -2.4% to -0.4%), Mauritius (mean *CFR*_*diff*_ = -3.3%; 95% HPD intervals -6.6% to -0.3%), Senegal (mean *CFR*_*diff*_ = -0.9%; 95% HPD intervals -1.4% to -0.4%), and Sierra Leone (mean *CFR*_*diff*_ = -3.8%; 95% HPD intervals -6.4% to -1.3%) (Figure 3 & Table S3).

**Figure 3:**
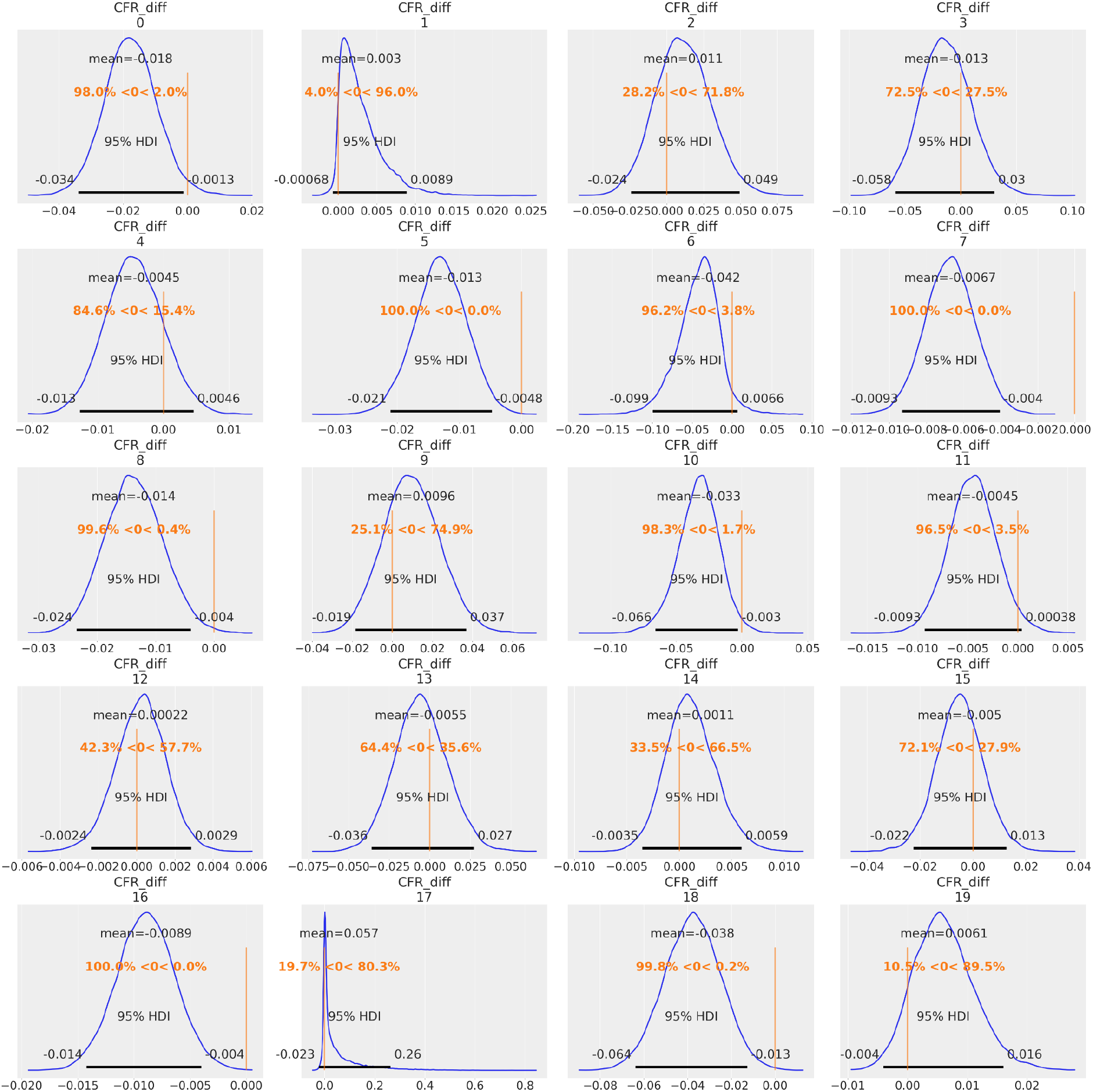
*Country-specific estimates, all age-groups combined:* posterior distributions of *CFR*_*diff*_ (= CFR in women - CFR in men), for each country. The horizontal black line represents the 95% HPDs. 95% HPD intervals are the same as that of 95% HDIs (= highest density intervals). 95% HPDs for 0: Angola (−0.034, -0.001), 5: Eswatini (−0.021, -0.005), 7: Guinea (−0.009, -0.004), 8: Kenya (−0.024, -0.004), 10: Mauritius (−0.066, -0.003), 16: Senegal (−0.014, -0.004), 18: Sierra Leone (−0.064, -0.013) do not contain 0 (vertical orange line), indicating significant differences between sex-specific CFRs in these countries. Panels 0-19 represent the following: 0: Angola, 1: Botswana, 2: Burkina Faso, 3: Chad, 4: Congo, 5: Eswatini, 6: Gambia, 7: Guinea, 8: Kenya, 9: Liberia, 10: Mauritius, 11: Mozambique, 12: Namibia, 13: Niger, 14: Rwanda, 15: São Tomé e Principe, 16: Senegal, 17: Seychelles, 18: Sierra Leone, and 19: Uganda.

In the ‘60+’ age-group, CFRs were significantly higher in men in seven countries: Eswatini (mean *CFR*_*diff*_ = -9.4%; 95% HPD intervals -17.0% to -2.1%), Guinea (mean *CFR*_*diff*_ = -4.2%; 95% HPD intervals -6.3% to -2.2%), Kenya (mean *CFR*_*diff*_ = -10.3%; 95% HPD intervals -18.2% to -2.7%), Mauritius (mean *CFR*_*diff*_ = -15.4%; 95% HPD intervals -29.6% to -2.3%), São Tomé e Principe (mean *CFR*_*diff*_ = -6.8%; 95% HPD intervals -13.7% to -0.8%), Senegal (mean *CFR*_*diff*_ = -4.1%; 95% HPD intervals -6.1% to -2.0%), and Sierra Leone (mean *CFR*_*diff*_ = -16.3%; 95% HPD intervals -29.6% to -2.6%) (Figure 4 & Table S3).

**Figure 4:**
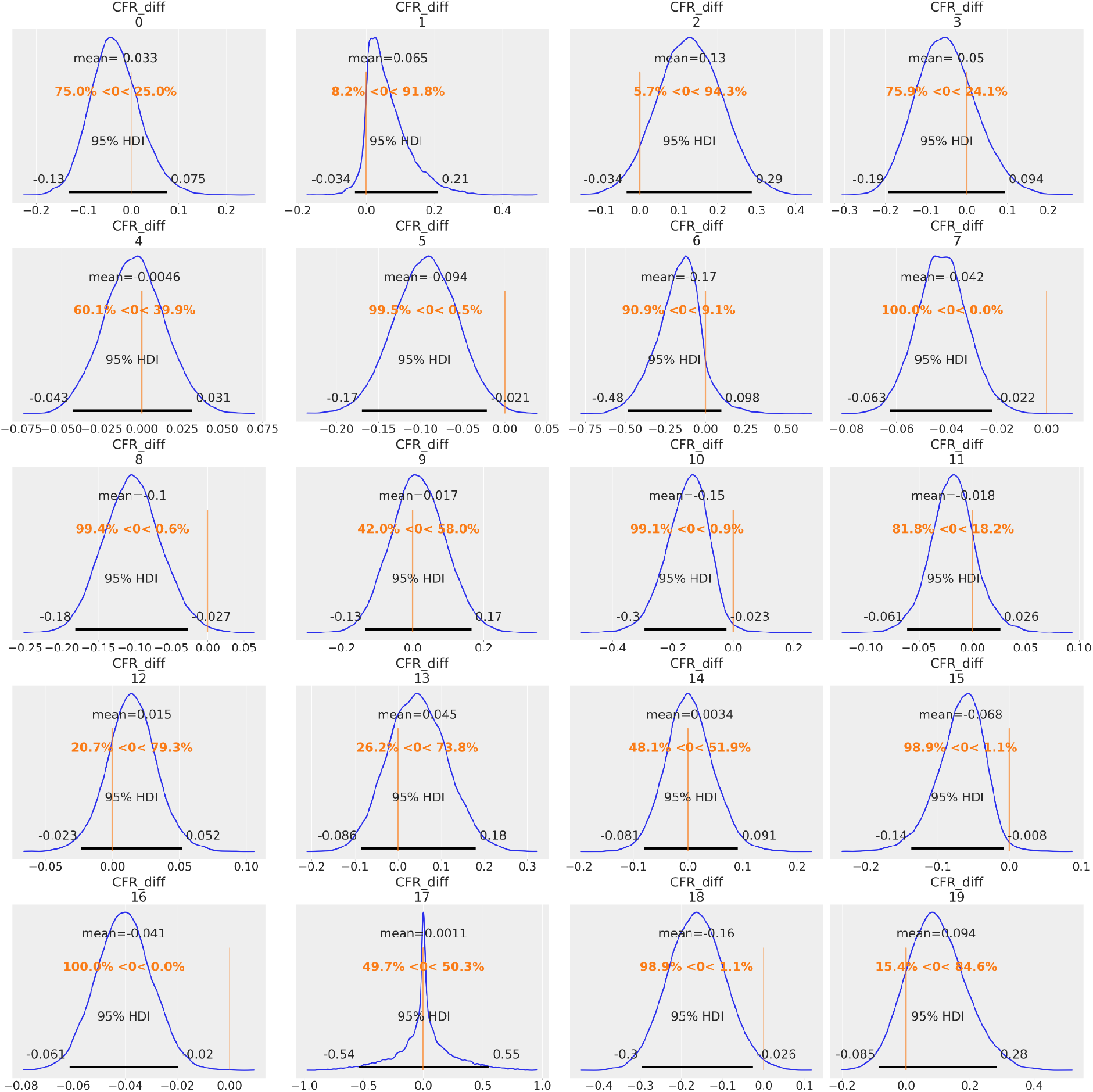
*Country-specific estimates, 60+ age-group:* posterior distributions of *CFR*_*diff*_ (= CFR in women - CFR in men), for each country. The horizontal black line represents the 95% HPDs. 95% HPD intervals are the same as that of 95% HDIs (= highest density intervals). 95% HPDs for 5: Eswatini (−0.17, -0.021), 7: Guinea (−0.063, -0.022), 8: Kenya (−0.182, -0.027), 10: Mauritius (−0.296, -0.023), 15: São Tomé e Principe (−0.137, -0.008), 16: Senegal (−0.061, -0.02), 18: Sierra Leone (−0.296, -0.026) do not contain 0 (vertical orange line), indicating significant differences between sex-specific CFRs in these countries. Panels 0-19 represent the following: 0: Angola, 1: Botswana, 2: Burkina Faso, 3: Chad, 4: Congo, 5: Eswatini, 6: Gambia, 7: Guinea, 8: Kenya, 9: Liberia, 10: Mauritius, 11: Mozambique, 12: Namibia, 13: Niger, 14: Rwanda, 15: São Tomé e Principe, 16: Senegal, 17: Seychelles, 18: Sierra Leone, and 19: Uganda.

In the ‘40-59’ age-group, Angola was the only country that had a significant CFR difference, with men having a higher CFR than women (mean *CFR*_*diff*_ = -2.5%; 95% HPD intervals -4.6% to -0.7%; Figure S4 & Table S3). No country demonstrated significant sex-specific CFR difference in the ‘0-39’ age-group (Figure S3 & Table S3).

#### Sensitivity analysis

Our sensitivity analysis using other priors (Bayes-Laplace and Jeffreys) yielded similar estimates as that of Kerman’s prior. Exceptions included countries with scarce data, for example, Seychelles, Botswana, and Gambia (Figure S1).

## Discussion

This study provided information on the sex and age specific difference in SARS-CoV-2 case fatality rate among 20 countries in the SSA until September 1, 2020. As one of the largest studies analysing CFR differences between women and men in SSA, our results showed that age-specific CFRs were higher among men than women in the SSA and that mean CFR differences increased with age.

Our data illustrates that at the regional sub-Saharan level men demonstrate higher crude CFR than women. Our SSA-specific estimates align with most epidemiological global studies where men were reported to have higher mortality than women ^6,8,18,19^. Moreover, several papers ^20,21^ showed that being a man places individuals at greater risk for health complications and death. The reasons for the gender difference in the fatality of COVID-19 could be attributed to sex-based biological factors and social, behavioral, or lifestyle factors related to gender. Several biological determinants like the immune system, genetics, sex hormones and the microbiome could contribute to lower COVID-19 fatality rates among women ^22,23^. Biological aspects are influenced by gender roles, norms, practices, and masculinities, which in turn, further affect health. For example, greater predispositions to health-harming behaviours among men (i.e. smoking ^24^) contributes to the development of non-communicable diseases and health complications later on in life ^25^. These health complications could lead to increased comorbidity-related mortality among men when paired with COVID-19 infection. Nevertheless, sex- and age-disaggregated data only explain a small fraction of the complex intersectional biological and social health inequities ^11^ that were exposed or have arisen from the COVID-19 outbreak. In order to comprehensively address gender-related disparities, one must also consider the intersecting links of social class, economic conditions, ethnicity, religion, and able-bodidness to sex and gender ^26^.

In contrast to such global evidence on the association between sex and COVID-19 CFR, some countries such as India (as of May 2020) ^27^, Nepal, Vietnam, Slovenia (as of September 18, 2020) ^28^ reported lower COVID-19 fatality rates in men than in women. Such differential findings might reflect incomplete COVID-19 data across countries, biases in case identification by sex, gender inequality in accessing healthcare facilities, or higher infection risks for women in certain countries. Akter Sonia ^29^ found that gender inequality in healthcare access, combined with limited health systems capacity, is likely to increase underreporting of COVID-19 fatalities among women in the United States (as of July 25, 2020). Gender biases in healthcare lead to incorrect diagnoses and poor treatment for both women and men, subsequently leading to worse health outcomes overall ^30^. Comprehensively understanding gender biases in healthcare settings are thus needed.

At the country level however, only seven out of 20 countries demonstrated a marked difference in mortality. A descriptive study from Niger that used sex-disaggregated individual-level data until July 2020 also found no statistical significance in CFR difference between men and women ^31^. Out of the 13 countries that did not demonstrate statistical significant CFR differences between women and men, three of them (Botswana, Seychelles, and Gambia) had low sample sizes; no statistical difference for these country-level samples could thus be attributed to lack of data variance within small samples. Further studies need to be conducted to confirm our findings.

Elderly populations may be disproportionately affected by COVID-19 owing to fragility due to aging and physiological changes, weaker immunity compared to younger people, and the increasing frequency of non-communicable comorbidities associated with age ^20,32,33^. Underlying conditions and comorbidities, such as hypertension, diabetes and cardiovascular disease, have been found to further exacerbate COVID-19 disease progression and fatality ^34,35^. Chronic diseases and comorbidities may themselves be gendered, generated through gendered behavioural and social pressures or expectations ^36^. Our findings reinforce that of global trends ^6,7,37^. While our study support global trends during this time period, further research is necessary and encouraged to understand the interaction between sex and age, particularly in a SSA context. Further research is necessary and encouraged to understand the interaction between sex and age, particularly in a SSA context. Prevention activities should target men and women over the age of 40.

### Strengths and limitations of the study

Thus far, to the best of our knowledge, this is one of the largest studies exploring sex-specific differences in CFRs at regional and country level of SSA. Our data was derived from individual-level patient data, which included information on age and sex, for a consecutive duration of 120 days from 20 SSA countries. Due to data unavailability, we were only able to include 20 from the 47 member states comprising the WHO African region. A particular strength of our study included our choice of prior. Our sensitivity analysis indicated that our prior was robust, since the results from all priors considered produced similar results. Among the 20 countries, only four countries had low sample sizes in some age categories, suggesting strong statistical validity and accuracy among the remaining 16 countries. Our study however, goes not without limitations. Our dataset was solely limited to age and sex; other gender-relevant indicators, such as presence of comorbidities, socioeconomic status, and immunological conditions were not available for analysis, preventing us from conducting further analyses on gender-related health inequities. Furthermore, gendered hospital admissions and differential treatment for men and women in clinical settings could act as a confounding source of bias to COVID-19 case and death reporting, impacting our CFR results. Due to lack of data, we could not address this. Moreover, case definition, surveillance capacity (e.g., variation in testing rates) and data management varied from country to country, which made comparisons across countries complicated.

Our results suggest that both sex and age are important predictors of COVID-19 mortality. Sex-disaggregated data and analyses are imperative to identity target risk groups, and thus reduce differences in COVID-19 exposure and vulnerability for both women and men in SSA ^38^. In addition to the collection of sex-disaggregated data on COVID-19 prevalence and mortality, disaggregated data on testing rates, hospitalisations, and healthcare workers provide further insights into COVID-19 sex differences, and should not be overlooked in future studies. Although not analysed in our study, it is important to emphasise that age and sex do not act alone, but intersect with other social determinants of health to influence COVID-19 disease progression and mortality. In order to design and promote gender-sensitive public health interventions it is essential that SSA countries collect and report up-to-date COVID-19 statistics that are disaggregated not only by sex and age, but also other social categories, such as class, economic status, and ethnicity ^11^.

## Data Availability

The data (data dictionary, statistical code) that support the findings of this study are available upon reasonable request from the corresponding author. Some of the data are publicly available through situation reports produced by Ministries of Health and WHO/AFRO on their respective websites.

## Contributions

JD and IT conceived the study, with the support of GGD. JD, IT, GGD, AJ, BN, and AV designed the study. JD and BN acquired, cleaned, and analysed the data. JD performed the statistical analysis with the supervision and support of FCC and AJ. JD, AJ, IT interpreted the data. JD, IT, AJ, GGD, BN, AV, SBM, and NL participated in weekly discussions that were helpful in exchanging ideas and making progress. IT, GGD and NL conducted the literature search. JD, IT, AJ, GGD, AV, BN drafted the initial manuscript, which was improved and finalized by JD, IT, GGD, AJ. FCC, BI, FM, GT, OK, CS, and JLA critically revised the manuscript for important intellectual content and supervised the study. All authors contributed to final approval of the version to be submitted. The corresponding author attests that all listed authors meet authorship criteria and that no others meeting the criteria have been omitted. JD, IT, FCC, and OK are the guarantors.

## Acknowledgements

We would like to express our sincere appreciation and gratitude to the entire team at the GRAPH Network and the WHO Regional Office for Africa for guiding and supporting us along the entirety of the study. Additionally, we would like to thank the governments of Angola, Botswana, Burkina Faso, Chad, Congo, Eswatini, Gambia, Guinea, Kenya, Liberia, Mauritius, Mozambique, Namibia, Niger, Rwanda, São Tomé e Principe, Sierra Leone, Senegal, Seychelles, and Uganda for their contribution to the merge line listing dataset.

## Ethical Approval

Data were collected for surveillance purposes under IHR, and no ethical approval was needed.

## Transparency statement

JD, IT, FCC, and OK affirm that the manuscript is an honest, accurate, and transparent account of the study being reported; that no important aspects of the study have been omitted; and that any discrepancies from the study as originally planned (and, if relevant, registered) have been explained.

