## Supplementary Information for "COVID-19 Mortality in Women and Men in Sub-Saharan Africa: A Cross Sectional Study"

**Table S1:** *SSA-specific estimates* (after merging the data from 20 SSA countries): mean estimates along with the 95% HPDs corresponding to the posterior distribution of CFR in women, CFR in men, and $CFR_{diff}$ ( = CFR in women - CFR in men), for each age-group and overall (all age-groups combined). We have highlighted the statistically significant differences in sex-specific CFRs using the bold font. Estimates are presented in percentages.

| **Age Group** | **CFR** | | $CFR_{diff}$ |
|---|---|---|---|
| | Women | Men | |
| **0-39** | 0.5 (0.4 to 0.6) | 0.6 (0.5 to 0.7) | -0.1 (-0.2 to 0.1) |
| **40-59** | 2.3 (2.0 to 2.7) | 3 (2.7 to 3.2) | **-0.7 (-1.10 to -0.20)** |
| **60+** | 9.4 (8.30 to 10.5) | 13.3 (12.4 to 14.30) | **-3.9 (-5.3 to -2.4)** |
| **Overall** | 1.8 (1.7 to 2) | 2.7 (2.6 to 2.9) | **-0.9 (-1.10 to -0.6)** |

**Table S2:** *Country-specific estimates:* mean estimates corresponding to the posterior distribution of CFR in women and CFR in men, for each age-group and overall (all age-groups combined).

| *Country* | | *CFR in women* | | | | *CFR in men* | | |
|---|---|---|---|---|---|---|---|---|
| *Age-group:* | *0-39* | *40-59* | *60+* | *Overall* | *0-39* | *40-59* | *60+* | *Overall* |
| Angola | 0.009 | 0.002 | 0.093 | 0.014 | 0.014 | 0.028 | 0.126 | 0.032 |
| Botswana | 0.001 | 0.003 | 0.075 | 0.003 | 0 | 0.001 | 0.011 | 0 |
| Burkina Faso | 0.023 | 0.036 | 0.384 | 0.082 | 0.015 | 0.069 | 0.255 | 0.071 |
| Chad | 0.039 | 0.116 | 0.21 | 0.092 | 0.037 | 0.111 | 0.26 | 0.105 |
| Congo | 0.014 | 0.038 | 0.134 | 0.040 | 0.012 | 0.04 | 0.139 | 0.044 |
| Eswatini | 0.002 | 0.018 | 0.098 | 0.013 | 0.003 | 0.031 | 0.192 | 0.026 |
| Gambia | 0.011 | 0.035 | 0.05 | 0.007 | 0.007 | 0.071 | 0.217 | 0.050 |
| Guinea | 0.001 | 0.002 | 0.013 | 0.002 | 0.002 | 0.007 | 0.055 | 0.009 |
| Kenya | 0.01 | 0.055 | 0.183 | 0.034 | 0.012 | 0.061 | 0.286 | 0.047 |
| Liberia | 0.019 | 0.101 | 0.269 | 0.068 | 0.012 | 0.068 | 0.252 | 0.058 |
| Mauritius | 0.02 | 0.006 | 0.016 | 0.010 | 0.003 | 0.061 | 0.169 | 0.043 |
| Mozambique | 0.001 | 0.01 | 0.023 | 0.004 | 0.005 | 0.012 | 0.041 | 0.009 |
| Namibia | 0 | 0.005 | 0.037 | 0.003 | 0.001 | 0.004 | 0.022 | 0.003 |
| Niger | 0.027 | 0.052 | 0.294 | 0.073 | 0.009 | 0.064 | 0.248 | 0.078 |
| Rwanda | 0.001 | 0.007 | 0.083 | 0.006 | 0.001 | 0.006 | 0.080 | 0.005 |
| São Tomé e Principe | 0.016 | 0.021 | 0.007 | 0.015 | 0.015 | 0.008 | 0.075 | 0.020 |
| Senegal | 0.004 | 0.017 | 0.031 | 0.011 | 0.001 | 0.019 | 0.071 | 0.020 |
| Seychelles | 0.124 | 0.197 | 0.125 | 0.060 | 0.006 | 0.005 | 0.124 | 0.003 |
| Sierra Leone | 0.025 | 0.107 | 0.307 | 0.066 | 0.028 | 0.120 | 0.470 | 0.104 |
| Uganda | 0.004 | 0.029 | 0.248 | 0.017 | 0.004 | 0.013 | 0.153 | 0.011 |

**Table S3:** *Country-specific estimates:* mean estimates along with the 95% HPDs corresponding to the posterior distribution of $CFR_{diff}$ (= CFR in women - CFR in men), for each age-group and overall (all age-groups combined), for each country. We have highlighted the statistically significant sex-specific differences in CFRs using the bold font.

| | Country | Overall | 0-39 age-group | 40-59 age-group | 60+ age-group |
|---|---|---|---|---|---|
| 0 | Angola | **-0.018 (-0.034, -0.001)** | -0.005 (-0.021, 0.012) | **-0.025 (-0.046, -0.007)** | -0.033 (-0.132, 0.075) |
| 1 | Botswana | 0.003 (-0.001, 0.009) | 0.001 (-0.003, 0.005) | 0.003 (-0.004, 0.014) | 0.065 (-0.034, 0.211) |
| 2 | Burkina Faso | 0.011 (-0.024, 0.049) | 0.008 (-0.019, 0.037) | -0.033 (-0.082, 0.018) | 0.129 (-0.034, 0.288) |
| 3 | Chad | -0.013 (-0.058, 0.03) | 0.002 (-0.036, 0.048) | 0.005 (-0.077, 0.088) | -0.05 (-0.194, 0.094) |
| 4 | Congo | -0.004 (-0.013, 0.005) | 0.001 (-0.005, 0.009) | -0.002 (-0.016, 0.012) | -0.005 (-0.043, 0.031) |
| 5 | Eswatini | **-0.013 (-0.021, -0.005)** | -0.001 (-0.005, 0.003) | -0.013 (-0.029, 0.004) | **-0.094 (-0.17, -0.021)** |
| 6 | Gambia | -0.042 (-0.099, 0.007) | 0.004 (-0.039, 0.057) | -0.036 (-0.183, 0.114) | -0.167 (-0.485, 0.098) |
| 7 | Guinea | **-0.007 (-0.009, -0.004)** | -0.001 (-0.003, 0) | -0.005 (-0.01, 0) | **-0.042 (-0.063, -0.022)** |
| 8 | Kenya | **-0.014 (-0.024, -0.004)** | -0.002 (-0.008, 0.005) | -0.006 (-0.027, 0.018) | **-0.103 (-0.182, -0.027)** |
| 9 | Liberia | 0.01 (-0.019, 0.037) | 0.007 (-0.012 , 0.026) | 0.032 (-0.028, 0.093) | 0.017 (-0.133, 0.165) |
| 10 | Mauritius | **-0.033 (-0.066, -0.003)** | 0.017 (-0.008, 0.058) | -0.055 (-0.118 , 0) | **-0.154 (-0.296, -0.023)** |
| 11 | Mozambique | -0.005 (-0.009, 0) | -0.004 (-0.008, 0) | -0.002 (-0.016, 0.013) | -0.018 (-0.061, 0.026) |
| 12 | Namibia | 0 (-0.002, 0.003) | -0.001 (-0.003, 0.001) | 0.001 (-0.004, 0.007) | 0.015 (-0.023, 0.052) |
| 13 | Niger | -0.005 (-0.036, 0.027) | 0.018 (-0.004, 0.045) | -0.013 (-0.058, 0.038) | 0.045 (-0.086, 0.18) |
| 14 | Rwanda | 0.001 (-0.004, 0.006) | 0 (-0.002, 0.003) | 0.001 (-0.01, 0.013) | 0.003 (-0.081, 0.091) |
| 15 | São Tomé e Principe | -0.005 (-0.022, 0.013) | 0.001 (-0.023, 0.024) | 0.013 (-0.011, 0.041) | **-0.068 ( -0.137, -0.008)** |
| 16 | Senegal | **-0.009 (-0.014. -0.004)** | 0.002 (0, 0.005) | -0.003 (-0.013, 0.008) | **-0.041 (-0.061, -0.02)** |
| 17 | Seychelles | 0.057 (-0.023, 0.263) | 0.118 (-0.052, 0.508) | 0.192 (-0.04, 0.743) | 0.001 (-0.541, 0.552) |
| 18 | Sierra Leone | **-0.038 (-0.064, -0.013)** | -0.003 (-0.021, 0.016) | -0.013 (-0.075, 0.047) | **-0.163 (-0.296, -0.026)** |
| 19 | Uganda | 0.006 (-0.004, 0.016) | 0 (-0.006, 0.006) | 0.016 (-0.009, 0.046) | 0.094 (-0.085, 0.282) |

**Figure S1:** *Sensitivity analysis:* showing mean estimates of $CFR_{diff}$ (= CFR in women - CFR in men) in different age-groups (‘0-39’, ‘40-59’, ‘60+’) and overall (i.e., all age-groups combined), for all the 20 countries, using three different uninformative priors. Similar Bayesian average estimates were obtained using the 3 different priors. However, when data was scarce (Botswana, Gambia, Seychelles), Kerman’s prior (purple circles) gave the most reasonable mean estimates, and hence we are presenting the results obtained using this prior in the main text.

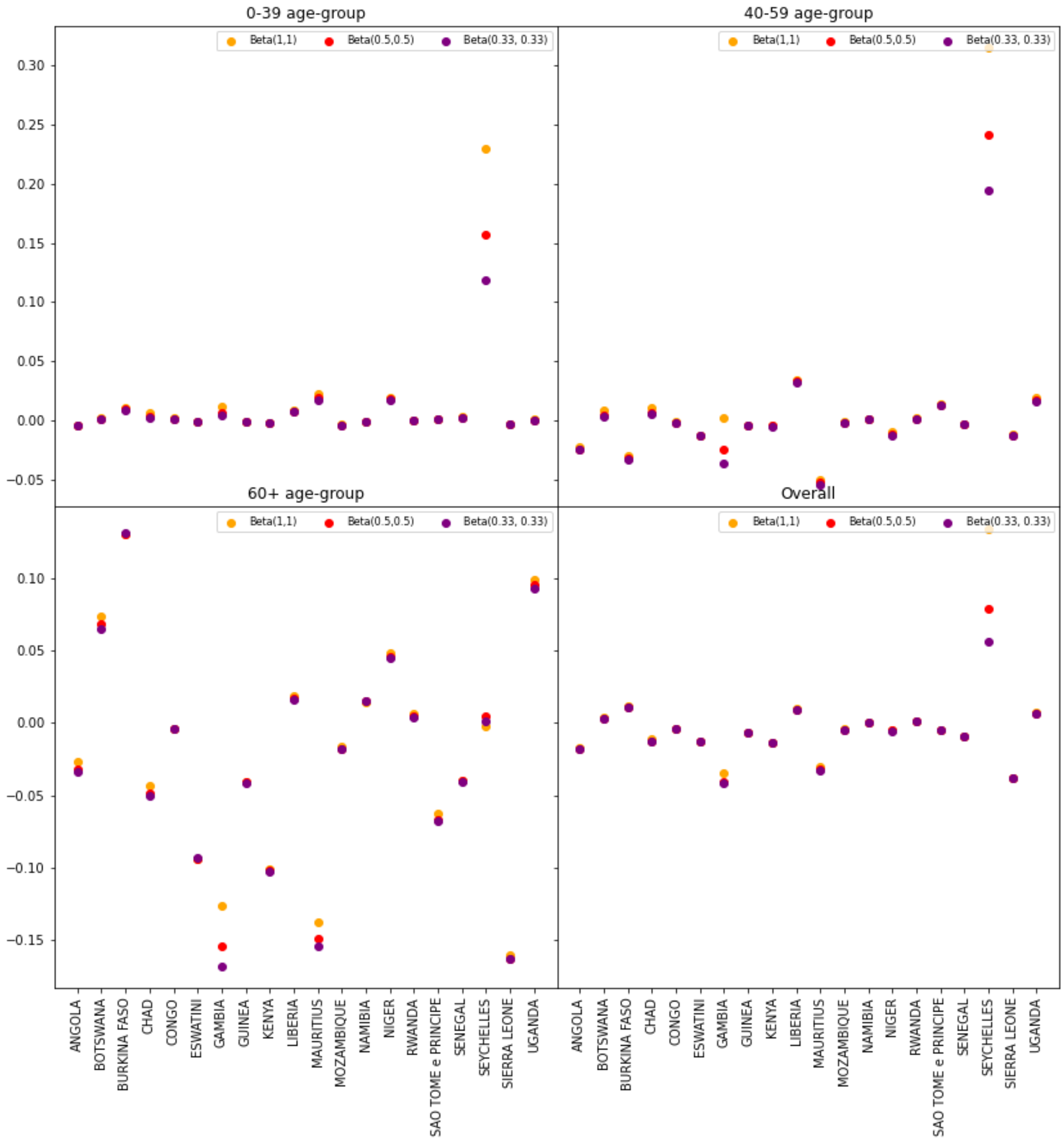

**Figure S2:** Age-specific CFRs for both sexes, for each country. Here, we are presenting the mean CFR estimates obtained using the Kerman's prior.

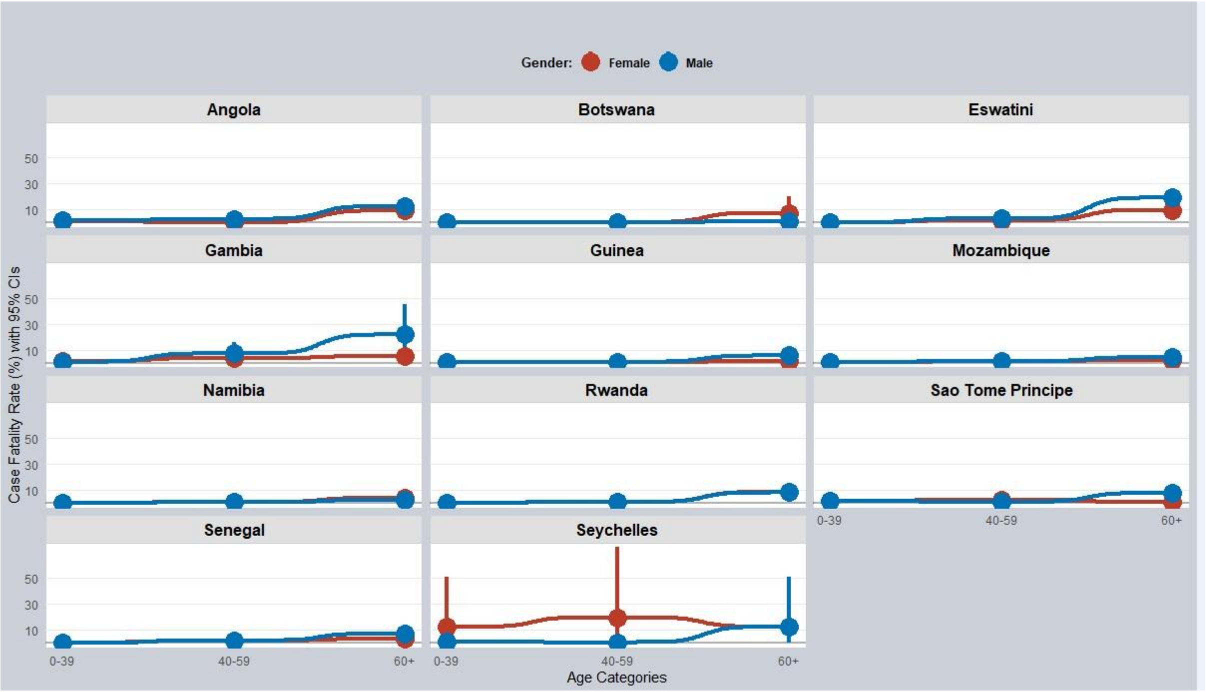


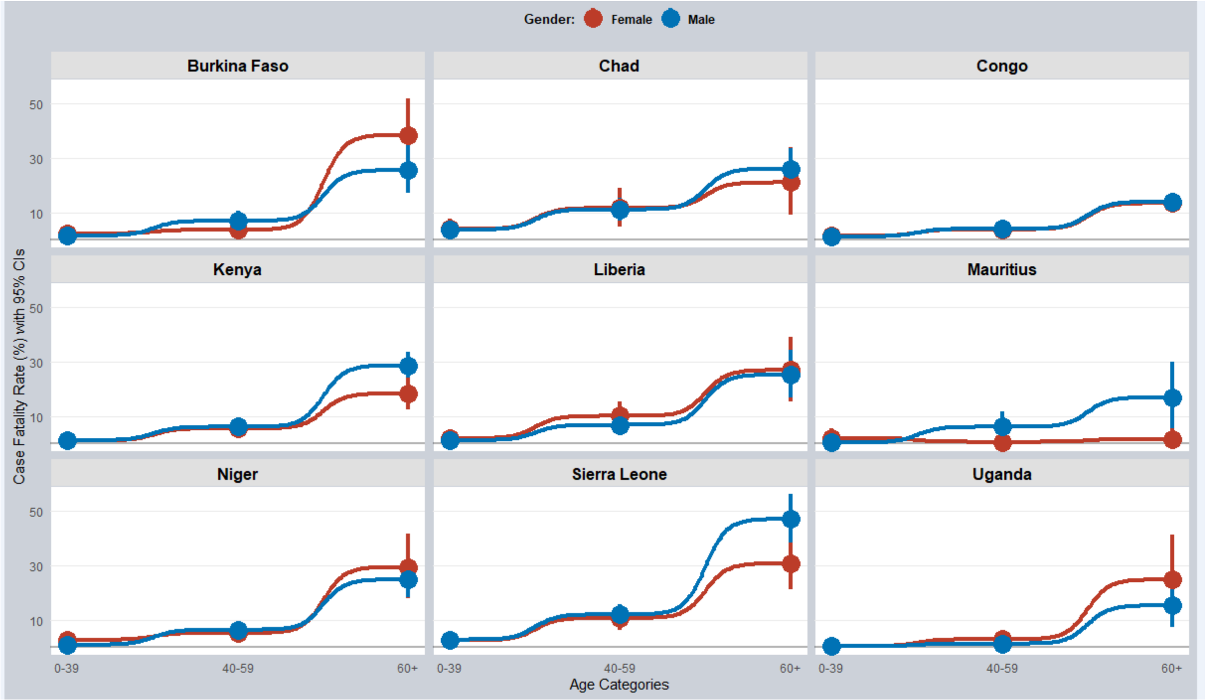

**Figure S3:** *Country-specific estimates, 0-39 age-group:* posterior distributions of $CFR_{diff}$ (= CFR in women - CFR in men), for each country. The horizontal black line represents the 95% HPDs. 95% HPDs are the same as that of 95% HDIs (= highest density intervals). 95% HPDs contain 0 (vertical orange line), indicating no significant differences between sex-specific CFRs, for all the countries. Panels 0-19 represent the following: 0: Angola, 1: Botswana, 2: Burkina Faso, 3: Chad, 4: Congo, 5: Eswatini, 6: Gambia, 7: Guinea, 8: Kenya, 9: Liberia, 10: Mauritius, 11: Mozambique, 12: Namibia, 13: Niger, 14: Rwanda, 15: São Tomé e Principe, 16: Senegal, 17: Seychelles, 18: Sierra Leone, and 19: Uganda.

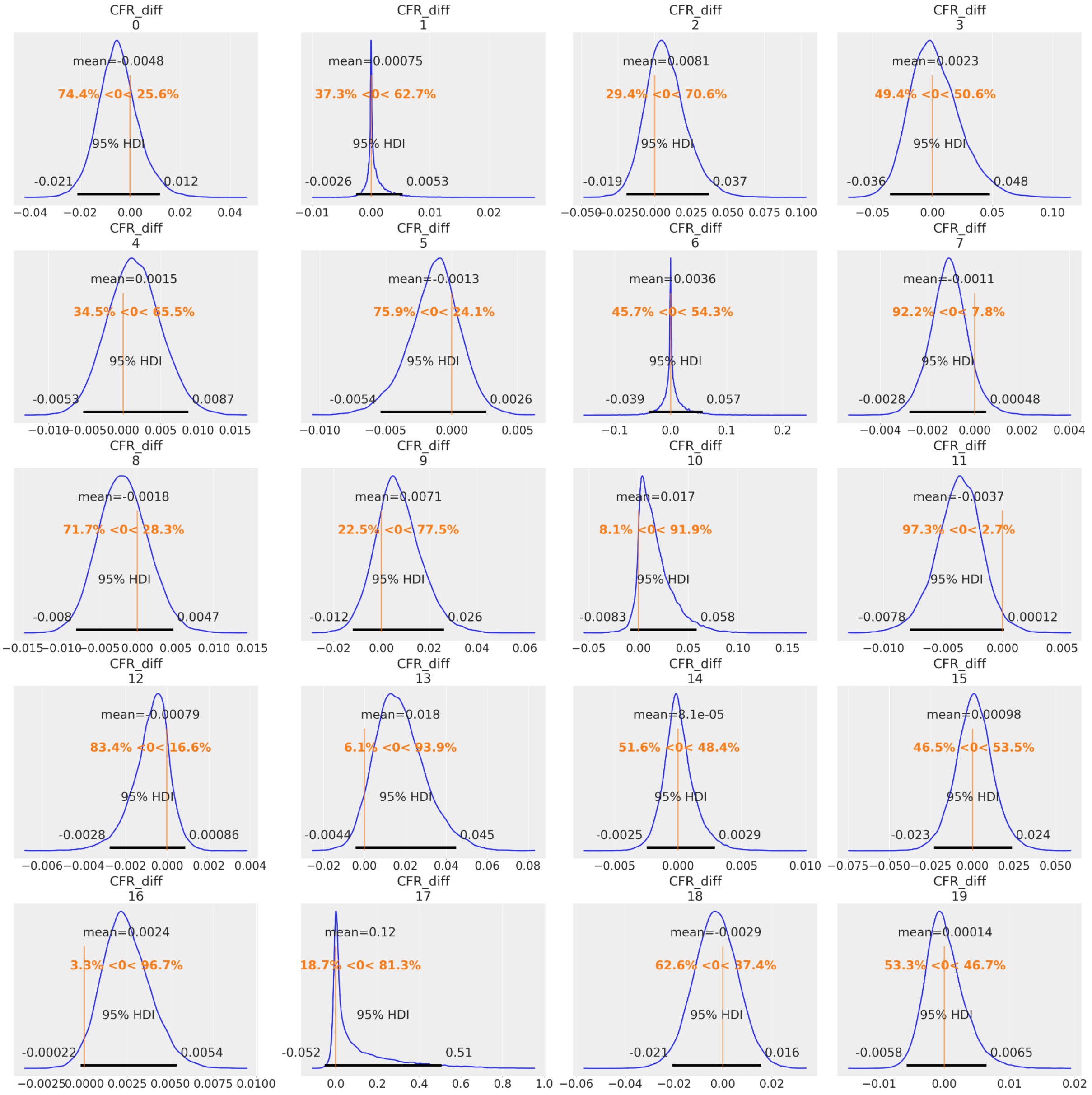

**Figure S4:** *Country-specific estimates, 40-59 age-group:* posterior distributions of $CFR_{diff}$ (= CFR in women - CFR in men), for each country. The horizontal black line represents the 95% HPDs. 95% HPDs are the same as that of 95% HDIs (= highest density intervals). 95% HPDs for 0: Angola (-0.046, -0.007), indicating significant differences between sex-specific CFRs in these countries. Panels 0-19 represent the following: 0: Angola, 1: Botswana, 2: Burkina Faso, 3: Chad, 4: Congo, 5: Eswatini, 6: Gambia, 7: Guinea, 8: Kenya, 9: Liberia, 10: Mauritius, 11: Mozambique, 12: Namibia, 13: Niger, 14: Rwanda, 15: São Tomé e Principe, 16: Senegal, 17: Seychelles, 18: Sierra Leone, and 19: Uganda.

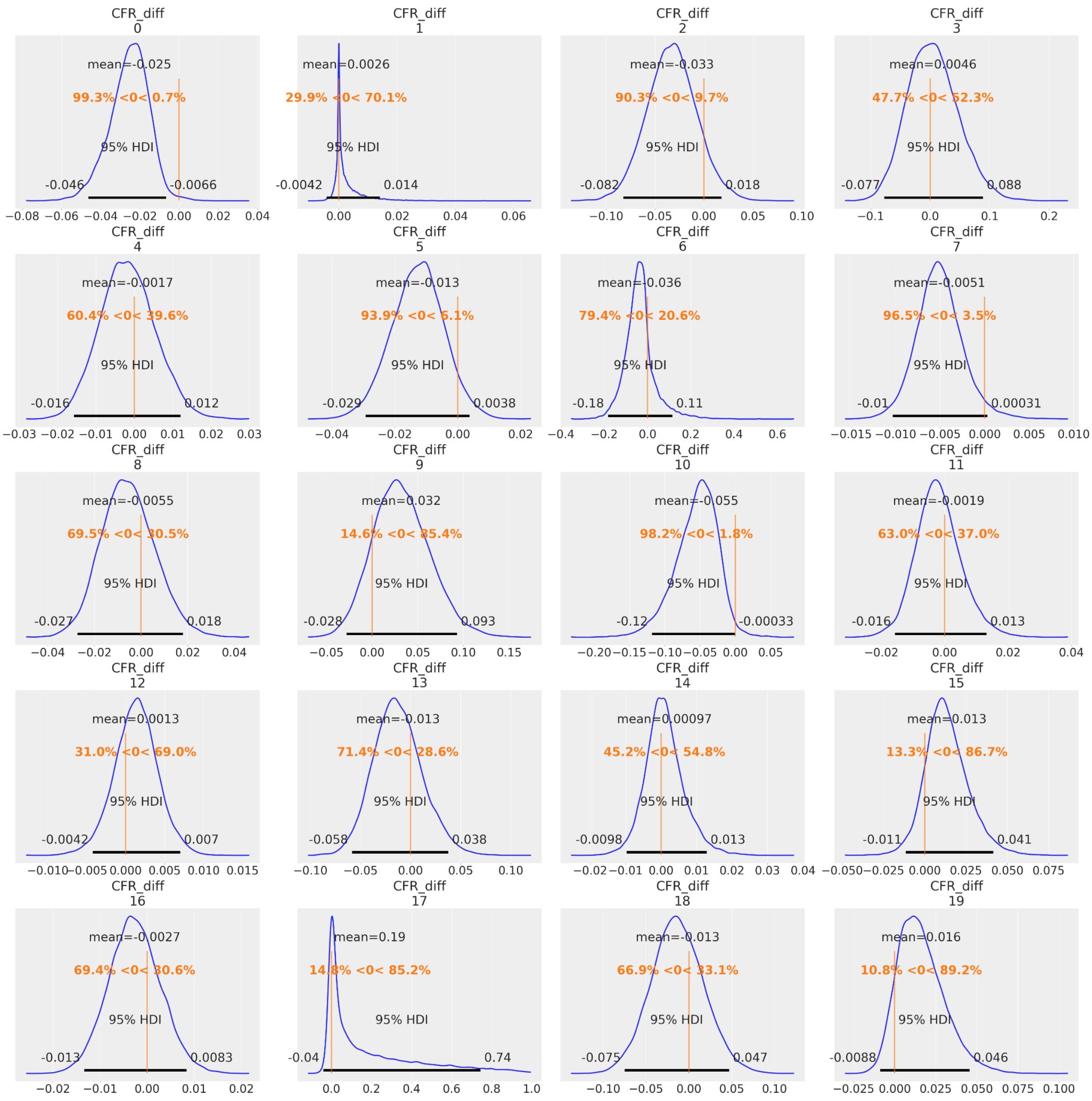
